# Aerosol emission of child voices during speaking, singing and shouting

**DOI:** 10.1101/2020.09.17.20196733

**Authors:** Dirk Mürbe, Martin Kriegel, Julia Lange, Lukas Schumann, Anne Hartmann, Mario Fleischer

**Affiliations:** Charité-Universitätsmedizin Berlin, Department of Audiology and Phoniatrics, Berlin, Germany; Technische Universität Berlin, Hermann-Rietschel-Institute, Berlin, Germany

## Abstract

Since the outbreak of the COVID-19 pandemic, singing activities for children and young people have been strictly regulated with far-reaching consequences for music education in schools and ensemble and choir singing in some places. This is also due to the fact, that there has been no reliable data available on aerosol emissions from children’s speaking, singing, and shouting. By utilizing a laser particle counter in cleanroom conditions we show, that children emit fewer aerosols during singing than what has been known so far for adults. In our data, the emission rates ranged from 16 P/s to 267 P/s for speaking, 141 P/s to 1240 P/s for singing, and 683 P/s to 4332 P/s for shouting. The data advocate an adaptation of existing risk management strategies and rules of conduct for groups of singing children, like gatherings in an educational context, e.g. singing lessons or choir rehearsals.

## Introduction

Aerosols are liquid or solid particles, which are transported in the air and not influenced by gravitation usually determined by a size less than 5 µm, that escape from the respiratory system during breathing, speaking and singing. Besides droplets, they are widely accepted carriers for the transmission of SARS-CoV-2 viruses (Morawska & Cao, 2020). Due to the principles of voice production and the described accumulation of SARS-CoV-2-infections during choir rehearsals (Hamner et al., 2020), it is assumed that singing is connected with increased aerosol emission rates. Recently, increased aerosol emissions during singing in comparison to speaking have been experimentally confirmed for adult singers (Alsved et al., 2020, Mürbe et al., 2020). Further, an increased aerosol emission rate is found for raising vocal loudness (Asadi et al., 2019). This results in limitations and specific risk management strategies especially for choir singing during the COVID-19-pandemic.

However, data about aerosol emission during singing for children are still missing. But especially for children and young people the restrictions on ensemble and choir singing have far-reaching consequences in addition to severe cultural and financial losses. Singing together is an obligatory part of school education and an important factor for the socio-emotional development of children and young people. This applies not only to music lessons in school, but also to the extracurricular sector with music schools and children and youth choirs. By now, the hygiene and performance concepts rely on aerosol emission rates during singing as collected from adults. For the first time, this pilot study presents data of aerosol formation when children sing.

## Results

Within the measuring range between 0.3 µm and 25 µm, about 99 % of all measured particles were smaller than 5 µm and more than 70 % smaller than 1 µm. With regard to the common understanding to denominate particles with a size smaller than 5 µm as aerosols the following results are cumulatively given for particles of size 0.3 µm – 5 µm.

The emission rates P_M_ for speaking were clearly lower than for singing (Figure 1). Whereas the median values for speaking were between 16 P/s (Particles/second) and 267 P/s, this measure was between 141 P/s and 1240 P/s for singing. For shouting, P_M_ was still higher with values from 683 P/s up to 4332 P/s. All subjects showed a clear individual increase in P_M_ for all three conditions. Linear mixed modeling showed, that these increases in condition were significant (likelihood-ratio test; p<.00001). On average, the ratio of P_M_ between singing and speaking was 5.87 ± 1.28 (standard error). For shouting and speaking, this ratio was 36.22 ± 1.28 (standard error). Both findings were significant (p<.001). Further, P_M_ was positively correlated with the maximum sound pressure level L_AFMAX_. An increase in one unit in L_AFMAX_ resulted in an increase in 0.05 units of log_10_ (P_M_). This finding was significant (p<.001).

**Figure 1.**
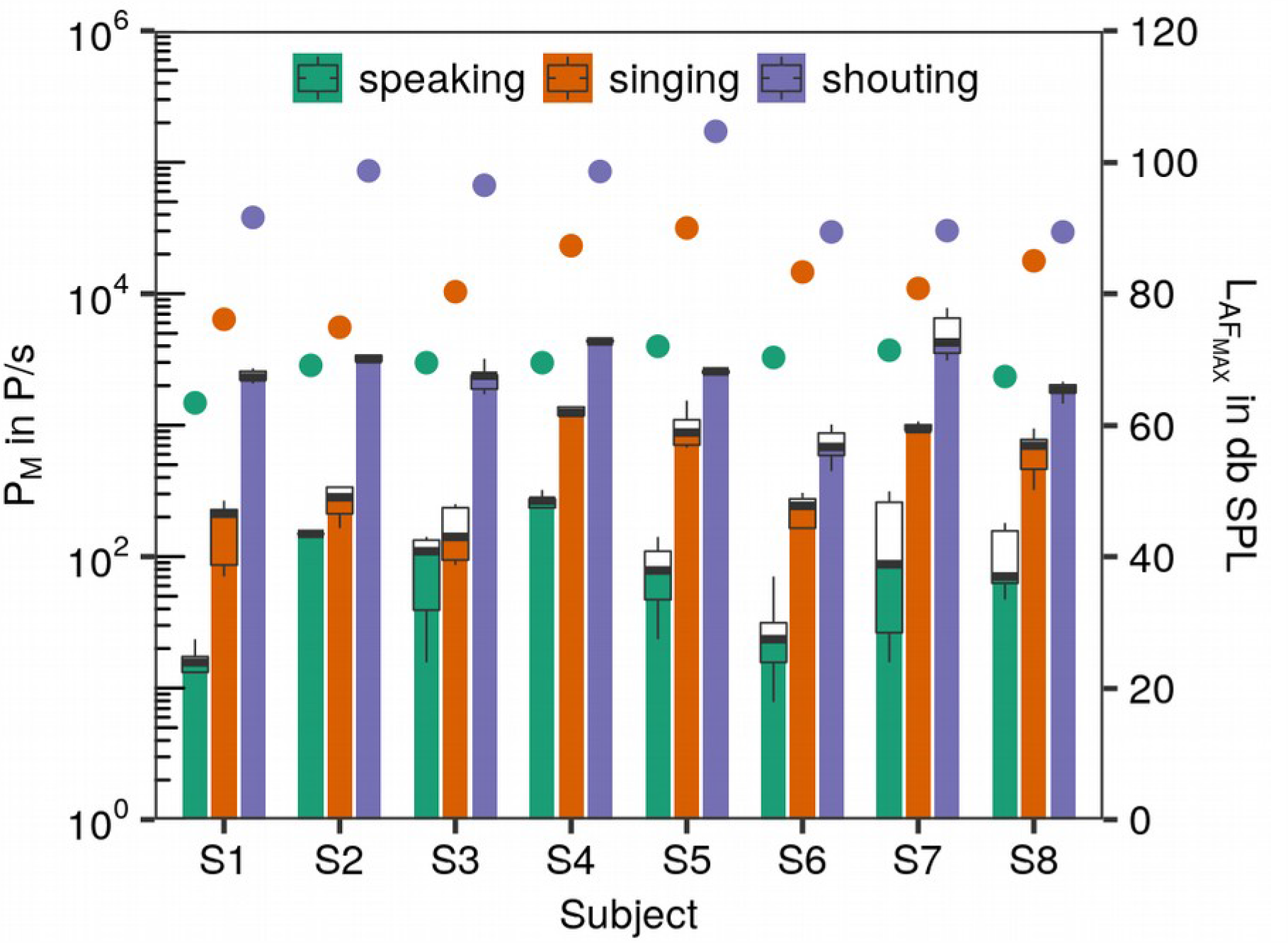
Boxplots of the emission rates (P_M_ in P/s, left y-axis) for the test conditions speaking, singing and shouting for subjects S1-S4 (girls) and S5-S8 (boys). The maximum sound pressure levels (L_AFMAX_ in db SPL) are also shown (right y-axis) with different colored full circles for the test conditions.

For the sustained phonation task, the median values for soft phonation (piano) were between 58 P/s and 683 P/s, this measure was between 58 P/s and 1907 P/s for loud phonation (forte). In contrast to the first task, not all subjects showed a clear increase in P_M_ from piano to forte. This finding was mirrored by the results of the linear mixed modeling approach. The increase of P_M_ from piano to forte was 1.91 ± 1.47 whereas condition was not significant (p=.133). Nevertheless, a positive correlation to L_AFMAX_ was found (Figure 2) which indicates that the emission rate increases with raising vocal loudness. Similar to the first task, an increase in one unit in L_AFMAX_ results in an increase in 0.05 units of log_10_ (P_M_) which was also significant (p<.001).

**Figure 2.**
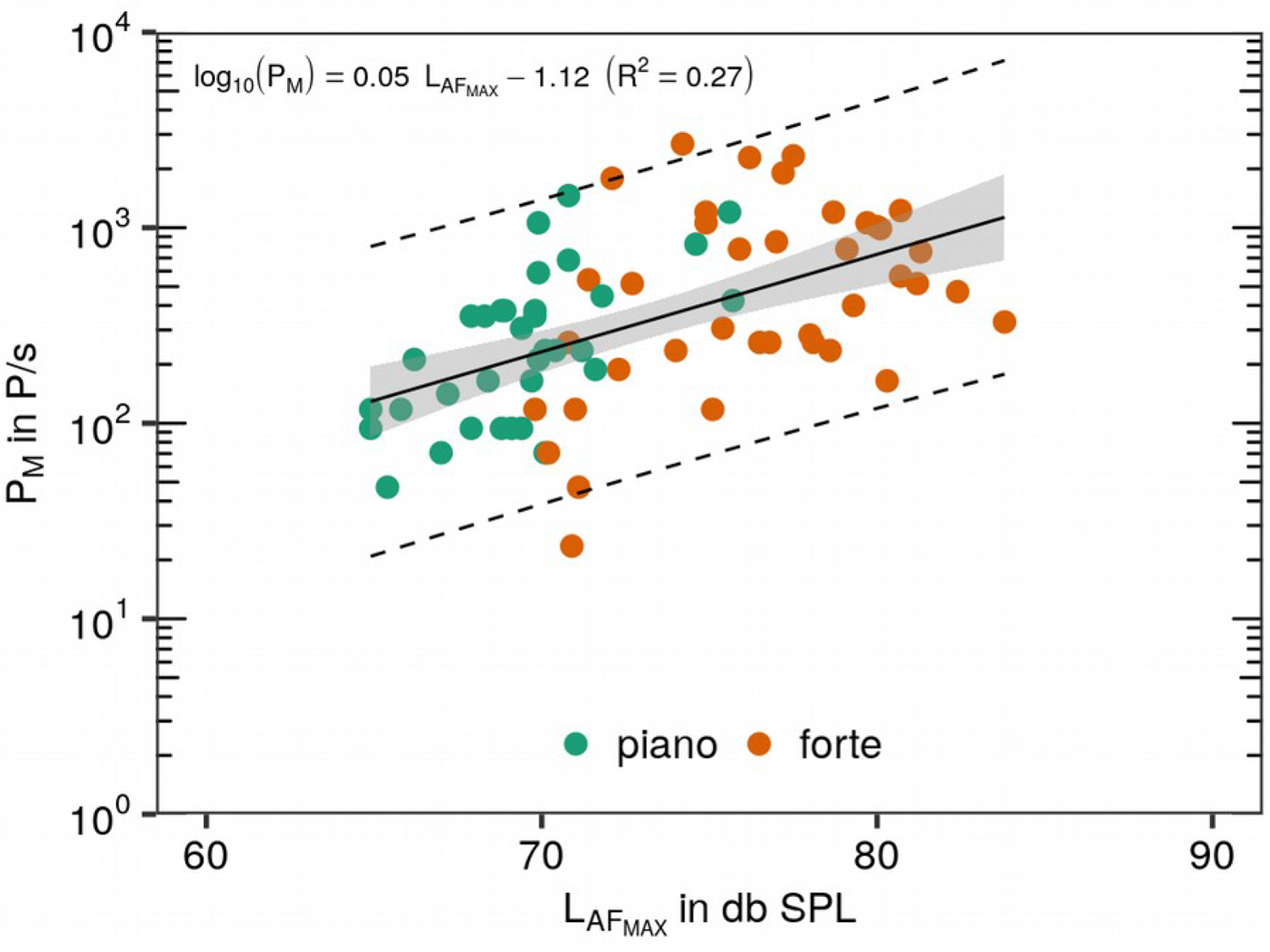
Emission rate P_M_ plotted over maximum sound pressure level L_AFMAX_ for sustained syllable /l*a*/. All five repetitions for the two loudness conditions are represented by colored points as denoted in the legend. The black solid line represents the linear regression (see inset for details), the gray colored area represents the 95% confidence region, whereas the black dashed lines restrict the 95% prediction band.

## Discussion

The present study confirms higher emission rates of aerosols for singing in comparison to speaking also for children. As for measurements of adult professional singers, a strong intersubject variability of aerosol emission was found for singing children, too. Finally, a positive correlation of particle emission with vocal loudness was confirmed, in particular reflected by the shouting condition.

Comparing these values with previously published data for adult professional singers (Mürbe et al., 2020, http://doi.org/10.5281/zenodo.4011701) similar values for speaking, but lower values for singing were observed (Figure 3). For singing, the ratio in medians between adults and children was about 3.1. Shouting values for children (not available for adults) were higher than singing values for adults. Regarding sustained phonation with loud voice, the ratio in medians between adults and children for the forte condition was about 6.8. It must be noted that there were slight deviations between adults and children in the execution of this task, such as children were allowed to shortly breathe within the recording sequence. Determined values for children are also lower than recently published data found in professional and non-professional adult singers (Alsved, 2020).

**Figure 3.**
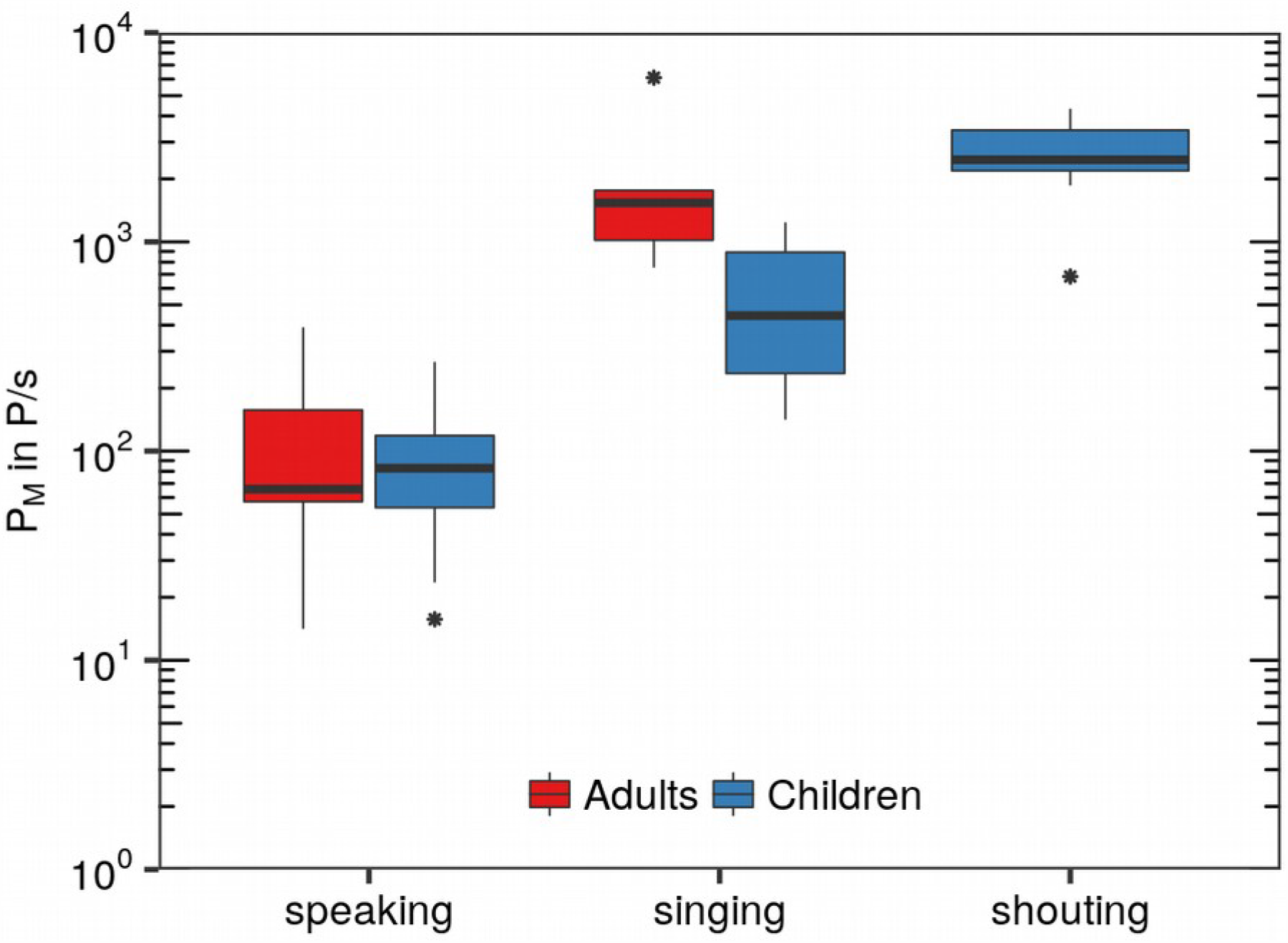
Boxplots of subject specific median emission rates (P_M_ in P/s) for the test conditions speaking, singing and shouting (children only). Present data of child voices (n=8) in comparison with previously measured corresponding data of professional adult singers (n=8) (https://doi.org/10.5281/zenodo.4011701, see Mürbe et al., 2020).

There might be different reasons for the lower emission rates for child voices during singing. Before mutation, there are considerable differences in the vibration characteristics of the vocal folds in comparison to adults. Typical features of a child’s vocal register in singing include differences in contact time and contact area of the vocal folds during each vibration cycle. There are also differences in the subglottic pressure between adults in general and children (McAllister & Sundberg, 1998, Howard, 2010). Further, there are smaller anatomical proportions of the child’s airways and vocal folds are shorter before mutation. On the other hand, the fundamental frequency of the voice and accordingly the contact frequency of the vocal folds might be higher, especially in comparison with male adult voices.

Indeed, a major reason for the lower emission rates might be the lower volume of the children’s voices during singing. This was especially evident in the task with intended loud singing, even if all children of this study had a longstanding choir experience. On the contrary, in the shouting condition, which is not related to limitations in the child’s singing technique, some children reached higher emission rates than adults during loud singing.

Our experimental setup uses a Laser Particle Counter (LPC) in cleanroom conditions (without a background concentration of particles) to assess the number and size of evaporated aerosols (or droplet nuclei) in their equilibrium state for different kinds of vocalisation. Estimating the precise number and size of these particles is of great interest to assess the concentration of those particles in closed rooms. Because of the great range of the exhaled air volume flow from zero (close mouth) up to at least 12-15 l/min for phonation (Gupta et al., 2010; Jiang et al., 2016; Mittal et al., 2013) and 24 l/min for blowing (Amis et al., 1999), the measurement setup must be both, highly sensitive to detect all particles and suitable to cover the whole volume flow range. Thus, different considerations were included in designing the setup for this study. First of all, a filter fan unit with a volume flow of 400 m^3^/h has been selected, whereas the flow of exhaled air is small compared to this and can be neglected. To avoid any disturbances regarding stagnated airflow at the measuring probe, this probe was positioned centrally in a glass pipe in which, caused by suction of the filter fan unit, the mean flow was only about 1.63 m/s. The resulting initial particle velocity during phonatory exercises was assumed to be 4.07 m/s in maximum and was added to the mean flow (see Mürbe et al., 2020 for details). Further, by placing a turbulence generating baffle between mouth opening and probe, there was a homogenous particle density distribution in the cross sectional area of the glass pipe. This in turn required the choice of an adequate distance between mouth opening and LPC, which was chosen to about 0.81 m and resulted in a traveling time of the particles of about 0.14 s in maximum. These experimental conditions, including a relative humidity of about 40%, result in approximately evaporated aerosols in their equilibrium state (Wei et al., 2015), which can be surveyed by the LPC with high accuracy and independently of the fluid flow at the mouth. Thus, the measured emission rates can serve as a realistic estimate for a possible carriage for viruses that propagate in the environment. Moreover, they allow a reliable comparison between the different vocalisation tasks. However, the emission rates reported in this study should not be interpreted as emitted droplets and aerosols directly at one’s opened mouth. Further issues with relevance for SARS-CoV-2–transmission during singing like the trajectory of larger droplets after emission from the mouth need to be studied with different methods like Particle Image Velocimetry or Phase Doppler Anemometry.

For the assessment of the risk of SARS-CoV-2-transmission during singing, both, droplets and aerosols are considered as virus carriers. While virus transmission via droplets can be mainly handled by distance and hygiene rules, the risk management of transmission through virus carrying aerosols has to be addressed with further strategies (Hartmann et al., 2020, Buonanno et al., 2020). Additional activities for safer ensemble and choir singing in closed rooms include limiting the number of singers and the rehearsal or performance time. This reduction of aerosol input leads to a lower cumulative aerosol concentration. On the other hand, room size and air condition systems will affect the number of potentially infectious aerosols in the room, too. Especially modern mechanical ventilation systems might significantly lower the risk of aerogenic virus transmission. Apart from the number of sources and the duration of singing, the individual emission rate of the singers determines the aerosol input in closed rooms. Based on the current prevalence of the disease, an advanced risk management for singing together should combine the above-mentioned tools. The lower aerosol emissions for children’s voices during singing might contribute to more sophisticated risk management strategies for singing in music lessons in school. These findings should be especially used to specify rehearsal and performance schedules for children’s and adolescent’s choirs because of the significance of education and socio-emotional development for children and young people.

## Materials and Methods

Four girls and four boys, all 13 years old (except one girl aged 15 years), participated in the study. All of them were members of a semiprofessional children’s choir (Staats- und Domchor Berlin, Mädchenchor der Singakademie Berlin) and had perennial choir experience between five and nine years. All children were prior to puberty vocal changes. The study was conducted according to the ethical principles based on the WMA Declaration of Helsinki and was approved by the ethic committee of the Charité-Universitätsmedizin Berlin, Germany. Informed written consent was obtained from all subjects and their parents.

The investigations were carried out in a cleanroom at the Hermann Rietschel-Institute, Technische Universität Berlin. In this highly pure environment, the subjects wore cleanroom clothing and a headgear to further reduce the clothing’s particle emission. To perform the experiments, subjects sat in front of the test equipment, consisting of a glass pipe with a diameter of 295 mm, through which a constant airflow of approximately 400 m^3^/h was generated by suction of a filter fan unit (Ziehl-Abegg, Künzelsau, Deutschland). The sampling probe (Ø37 mm) of a laser particle counter (LPC) (Lighthouse Solair 3100 E, Lighthouse Worldwide Solutions, Fremont, CA) was placed centrally in the pipe. The particle counter was counting with a sampling flow rate of 28.3 l/min with a measuring time increment of 10 seconds. The detected particles were assigned to six size classes between > 0.3 µm – 25 µm. The emission rate P_M_ was computed based on scaling of the particles measured at the LPC to the volume flow of the whole glass pipe. Apart from the particle measurement, the sound pressure level L_AFMAX_ was measured via a calibrated sound level meter (CENTER 322_ Datalogger Sound Level Meter, Center Technologies, Houston, TX). For detailed information about the cleanroom conditions, the particle measurement set-up and the audio equipment see Mürbe et al. (2020).

In the first task, the emission rates for three different vocal test conditions were compared: (a) speaking, (b) singing, and (c) shouting. Condition (a) was reading a standardized text (“Der Nordwind und die Sonne” by Äsop), (b) singing the Swedish folk song “Vem kan segla” in key G-Major. For condition (c), children were asked to cheer enthusiastically about a soccer game goal. The time window for a measured sequence was 30 seconds for test conditions (a) and (b) and 10 seconds for test condition (c). Each test condition was repeated five times.

In the second task, sustained phonation about 10 seconds was performed to investigate the impact of vocal loudness on the emission rate. Subjects were asked to sustain the syllable /la/, pitch G4 (392 Hz), at the two loudness conditions soft voice (piano) and loud voice (forte). To facilitate the 10 seconds measurement time, the children were allowed to take a short breath within the recording and to repeat the syllable.

Statistical analysis, individually handled for the two tasks, was carried out by using linear mixed model analysis in the statistical software R including the package lmerTest (Kuznetsova et al., 2017). Log-valued P_M_ data were incorporated as the response variable and condition as fixed effect. Further, intercepts for subject and by-subject random slopes for the effect of condition were regarded as random effects. P-values were obtained using Satterthwaite’s degree of freedom method.

## Data Availability

Publication of raw data are actually under preparation

## Acknowledgments

We thank the girls and boys and the accompanying staff of the Staats- und Domchor Berlin and the Mädchenchor der Singakademie Berlin for their personal support, and T. Nawka for general discussion.

## Author Contributions

D.M., M.F., and M.K. designed research. J.L., L.S., D.M. and M.F. made measurements. M.F. and D.M. analyzed data, M.F., D.M., J.L., L.S., A.H., and M.K. wrote the paper.

